# Infrared assessment of human facial temperature in the presence and absence of common cosmetics

**DOI:** 10.1101/2020.03.12.20034793

**Authors:** Kaikai Zheng, Ruoyu Dong, Huan Wang, Steve Granick

**Author notes:** these authors contributed equally (Kaikai Zheng, Ruoyu Dong, Huan Wang). Electronic mail (Steve Granick).

## Abstract

Using a sensitive research-grade infrared camera, we find that common facial cosmetics and lotions mask skin temperature in assays of the human forehead. We test a family of 10 commonly-used cosmetic products and find that volatile liquids and creams lower thermal skin temperature by at least 2 C for up to 5-10 min and at least 1 C for up to 20 min, respectively. Powder and cream that contains brightening agent lower indefinitely the skin temperature sensed by infrared camera. With the qualification that these experiments were performed in a controlled laboratory setting rather than the mass crowd screening environment where infrared temperature sensing of humans sees widespread use, our tests suggest that for human subjects whose face was treated with certain cosmetics and lotions, infrared-based screening for elevated facial temperature (fever) can be unreliable.

## I. INTRODUCTION

The average person trusts the validity of fever screening by noncontact infrared-based (IR) temperature measurement, which has become a staple of modern screening for elevated temperatures associated with coronavirus COVID-19 and other illness^1-8^. Such people with elevated temperature potentially should be denied access to sensitive locations such as a meeting room or hospital, or potentially should even be quarantined. It is interesting and relevant to consider limitations of the measurement on the basis of which such decisions are made, especially as the premise of this test is that skin temperature is an accurate proxy for body temperature.

This study assesses the hypothesis that skin temperature can be lowered simply by applying facial cosmetics. In this hypothesis, skin coating using creams, powders, sunscreen, and other instruments of personal care might lower the temperature measured by infrared imaging. Testing the hypothesis, we present here the results of screening a large family of 10 common cosmetic products.

## II. EXPERIMENTAL

For noncontact infrared thermal measurement^9,10^, we used a low-noise, high-performance mid-infrared research grade camera, FLIR A8303sc (FLIR, Inc., Wilsonville, OR, USA) with nominal resolution <0.02 ^*°*^C. The measurements were quantified using the company’s software, FLIR ResearchIR MAX. The measurements, made at 3 Hz, were averaged over 30 s to produce the data presented here.

The human subject of our tests was a male approximately 30 years old such that his skin lacked visible wrinkles. For quantification, we analyzed temperature on his forehead, as is typical for measurement scanning devices. One side of his forehead was used as a blank to which no cosmetic was applied, while the other side contained applied cosmetic. Temperature difference between these two spots was quantified. Usually the area of each spot was round, at least 2 cm in diameter.

Samples of cosmetics were selected from among those in common personal care use by researchers in our immediate environment. They were equilibrated at room temperature. Depending on the texture, they were applied in different ways. Toners and lotions were applied using common cosmetic cotton pads. Emulsions and creams were applied with hand. Powders were applied with makeup brush. Amounts similar to or slightly larger than those for daily use were applied.

## III. RESULTS

The 10 cosmetic products that we investigated are summarized in Table I.

**TABLE I.**
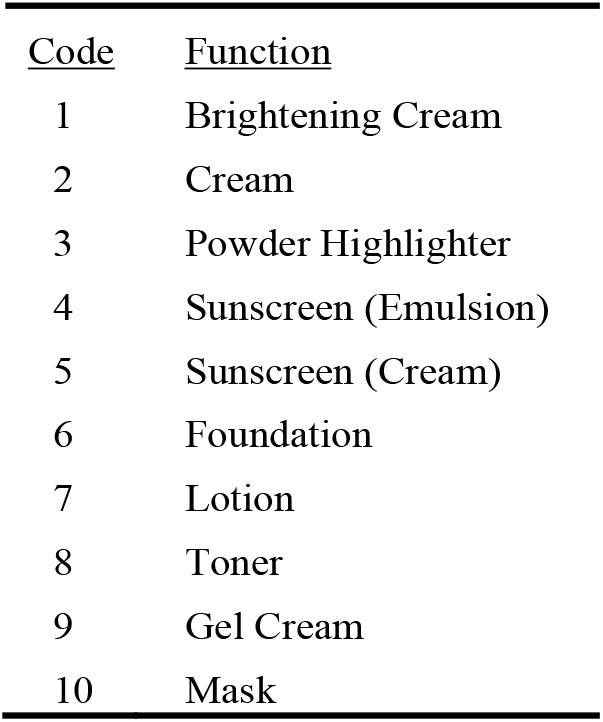
Family of 10 commercially-available cosmetics that were tested, identified by number-code and function. As the sources used are believed to be representative of the functions, brands are not identified.

### Raw data

*Fig. 1* illustrates raw data. The left panel (*Fig. 1a*), a typical thermal image, shows skin temperature measured in different regions of the subject’s face. One complication is that the subject inevitably shifted position of his head during the long measurement times, thereby presenting the possibility that the apparent temperature might shift owing to change in the measurement focus. To compensate for this possibility, we made differential measurements between two regions of the forehead, one blank region (cosmetic-free), one region to which cosmetics were applied. Showing the excellent reproducibility of the measurement over extended times, *Fig. 1b* illustrates apparent absolute temperature plotted against time. Naturally, this healthy subject’s skin temperature of ≈ 34.35 C was below his body temperature, which was normal.

**FIG. 1.**
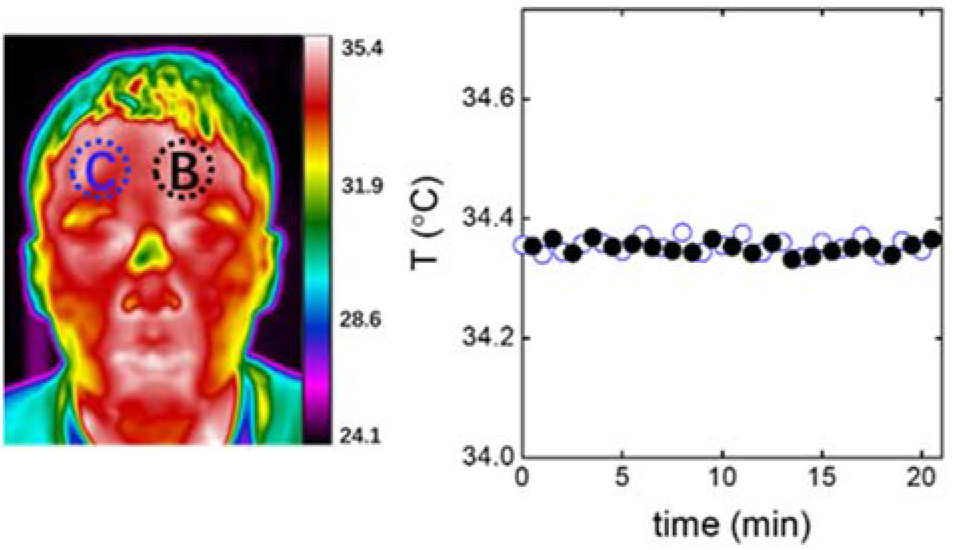
Raw data. Left panel: representative infrared image of the subject’s face. Data regarding temperature depression were quantified as the difference between the blank (“B”) and cosmetic-containing (“C”) regions of the forehead. The image includes a color bar identifying temperature. Right panel: Absolute skin temperature monitored in region C (open symbols) and B (filled symbols) versus time, showing excellent stability of the measurement.

### Creams, powder

In *Fig. 2*, the effect of applying cosmetic is considered. Depression of skin temperature is plotted against time after the cosmetic was applied. Cosmetic #1, marketed as a tone-up brightening cream, produced the largest depression of skin temperature. Skin temperature was depressed by at least 2 C for 15 min and 1 C for 20 min, likely because evaporation of volatile components lowers temperature. The evaporation rate was notably slow, perhaps because the volatile components (water) also dissolve in the skin. After the cosmetic had remained on the skin for 40 min, it was washed with cosmetic remover and strikingly, the skin temperature remained depressed by ≈ 0.8 C. This semi-permanent depression of skin temperature likely reflects the presence of residual solid-state brightener specified as an active ingredient in the product’s marketing.

**FIG. 2.**
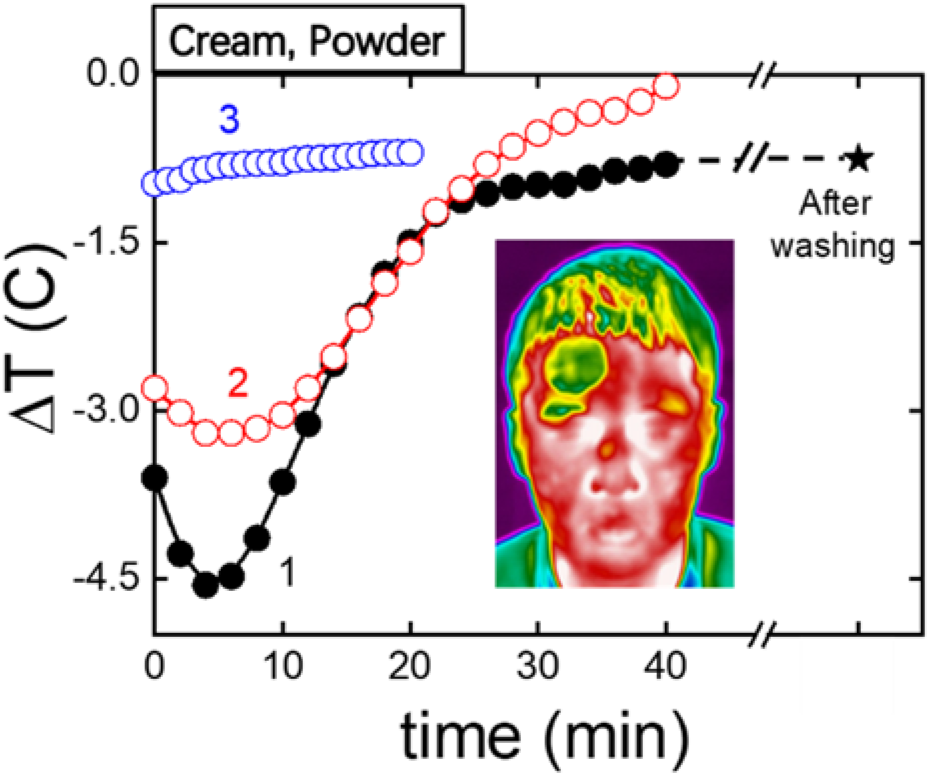
Cream and powder influence on thermal temperature measurement. Temperature depression is plotted against time for cosmetic #1, marketed as a tone-up brightening cream; cosmetic #2, marketed as a natural moisturizing cream; and cosmetic #3, a powder highlighter. The inset, a thermal image of the subject’s face, illustrates the cosmetic-coated region of the forehead.

Cosmetic #2, marketed as a natural moisturizing cream, also depressed the skin temperature by at least 2 C for 15 min and 1 C for 20 min. After this time, the skin temperature continued to decrease, probably reflecting the continuing absorption of cream into skin. After 40 min, no further change was observed.

Cosmetic #3, a powder highlighter containing dry powder, mica and talc, produced similar depression of skin temperature as the long-time effect of cosmetic #1, except that it lacked the strong temperature depression associated with evaporation of copious volatile liquid. The steady-state temperature depression was ≈ 0.8 C.

### Sunscreen

In *Fig. 3*, depression of skin temperature is plotted against time after sunscreens were applied. Cosmetic #4, an emulsion sunscreen, produced no depression of skin temperature after 5 min, reflecting rapid evaporation of the alcohol carrier liquid. However, cosmetic #5, a broad-spectrum cream sunscreen, produced markedly long-lived depression of the skin temperature for times extending beyond 30 min. Evidently these effects are not in proportion to the SPF (sun protection value) as SPF=50 and 30 for the former and latter, respectively. Skin temperature depression was less for the sunscreen with higher SPF. Instead, the remarkably strong temperature depression produced by cosmetic #5 appears to be that, just as for cosmetics #1 and #2, it is a cream.

**FIG. 3.**
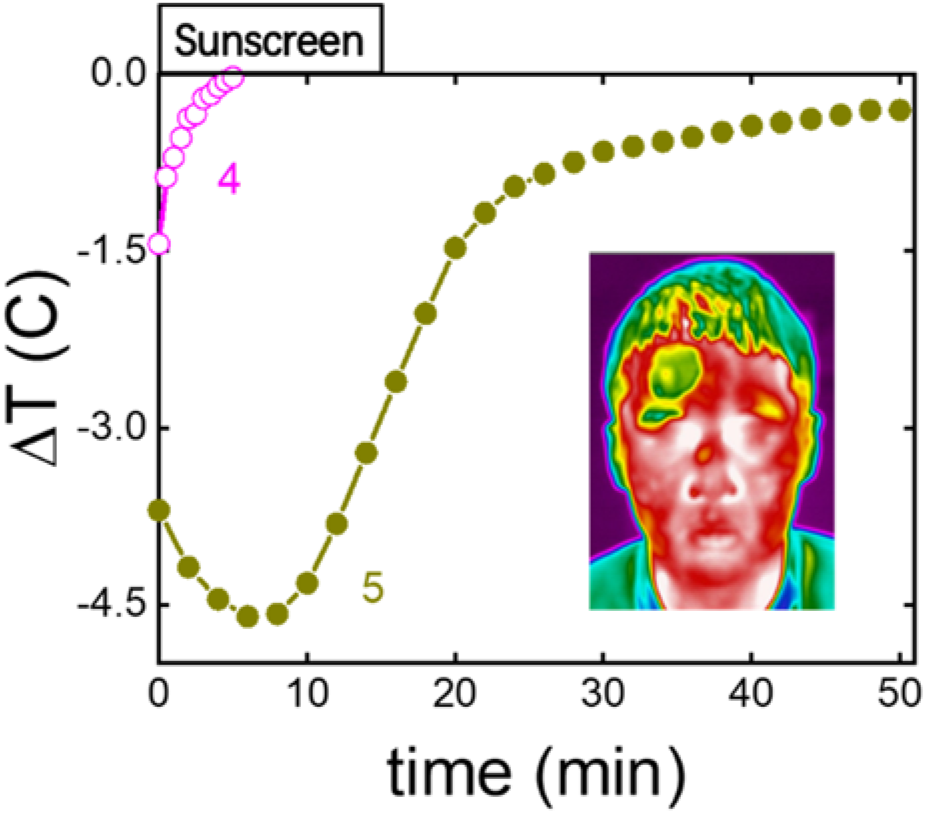
Sunscreen influence on thermal temperature measurement. Temperature depression is plotted against time for cosmetic #4, an emulsion sunscreen, and cosmetic #5, a broad-spectrum cream sunscreen. The remarkably strong temperature depression of cosmetic #5, despite its lower SPF value, appears to be because it is a cream. The inset, a thermal image of the subject’s face, illustrates the cosmetic-coated region of the forehead.

### Other cosmetics

In *Fig. 4*, depression of skin temperature is plotted against time for 5 additional cosmetics and for reference the strong temperature-depression effect of cosmetic #1 is included. Cosmetic #6, a foundation common in facial makeup, showed negligible effect after 5 min, despite the fact that the likely major components of residue are talc, mica, silica, dimethicone, zinc stearate, and titanium oxide. Cosmetic #7, a lotion with more volatile components, produced stronger temperature depression during the initial 5 min, but after 10 min there was no effect. For cosmetic #8, a toner, and cosmetic #9, a water-like gel cream, the effect likewise disappeared after 10 min.

**FIG. 4.**
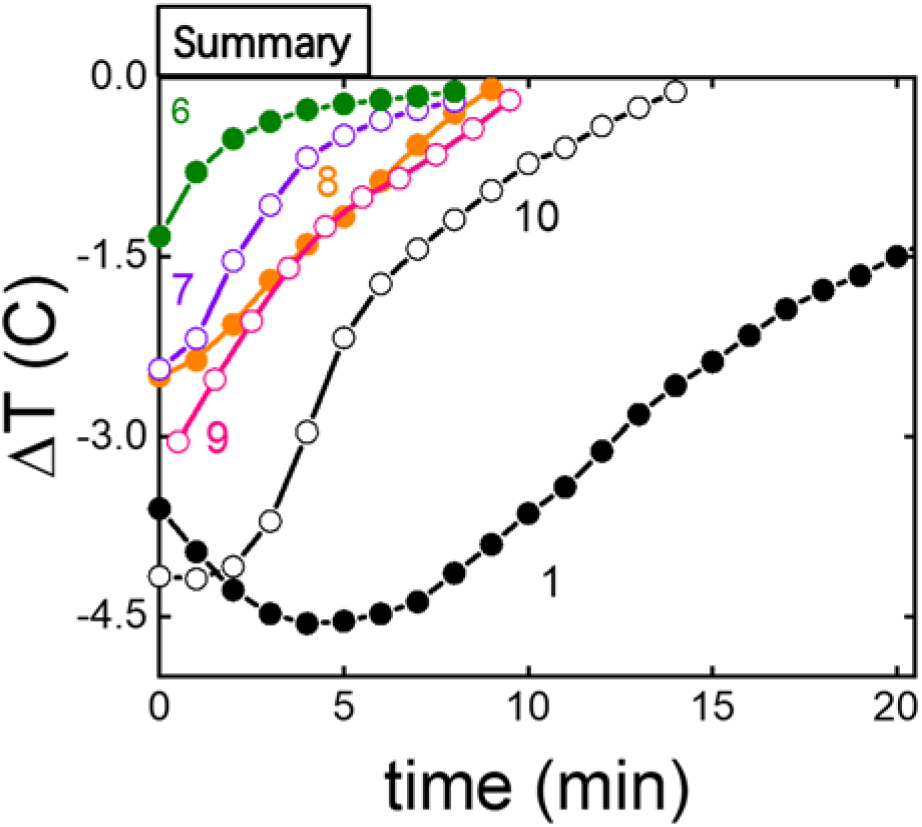
Other cosmetics. Depression of skin temperature is plotted against time for 5 additional cosmetics with the strong temperature-depression effect of cosmetic #1 included for reference: cosmetic #6, a color foundation common in facial makeup; cosmetic #7, a lotion with more volatile components; cosmetic #8, a toner; cosmetic #9, a water-like gel cream; and cosmetic #10, a facial mask used to moisturize the skin.

Cosmetic #10, a facial mask used to moisturize the skin, was left on the face for 30 min and skin temperature was monitored subsequently. A large effect was observed for the initial 10 min but there was no effect after 15 min.

## IV. CONCLUSION

The application of cosmetics to the human face can depress the temperature read in noncontact thermal imaging. The amount of depression falls into the following categories:

1. Cosmetics that lower skin temperature by the physical principle of evaporative cooling. The fact that liquids absorb heat to evaporate has been used for thousands of years for air conditioning, at least as long ago as in ancient Egypt, Persia, and India.^11^ In cosmetics, this short-term effect appears to disappear after 5-15 min.
2. Cosmetics that lower skin temperature by acting as a barrier layer between skin and surrounding air. The effect is marked for creams but the effect appears to disappear after 20-30 min, perhaps as cream is absorbed into the skin.
3. Cosmetics that lower skin temperature because they contain solids. The effect was most marked for brightening cream and powder highlighter, cosmetics #1 and #3, respectively. Depression of skin temperature appears to persist semi-indefinitely, probably because solid particles in the cosmetic, with lower emissivity than skin, are embedded in the skin. However, this effect is less prominent for foundation (cosmetic #6), suggesting that size of the solid particles matters.

The study will also apply to the reliability of portable temperature scanners which do not require imaging and pixel reading that we used here.^1-8^ However, it has no implication for infrared measurements of ear temperature^12^, as cosmetics are not applied to the eardrum.

It will be interesting and relevant, in future work, to understand how the ingredients of different cosmetic brands matters; in this study, we simply employed cosmetics that are in common use in our cultural environment. Also it will be interesting and relevant, in future work, to understand the influence of the amount of cosmetic that is applied; as noted above, we simply employed thickness roughly that which we find in common use around us. The purpose of this report is to call attention to the importance of the phenomenon and to show that cosmetics disguise actual skin temperature. Finally, in future work it will be interesting and relevant to understand better the physical reasons behind the phenomena reported above. Human skin has very high emissivity, 0.98, close to 1. Lower emissivity (higher reflectivity) will result in a lower value of temperature measured by the infrared (IR) camera. The IR camera does not measure true temperature, but actually measures radiated light from the object, which is then material and surface property dependent

These conclusions are important as they show that the temperature “seen” by infrared thermal screening can be easily disguised by common cosmetics. They are relevant to evaluating screening of human populations for the COVID-19 virus and other illnesses that cause fever. They suggest conclusions with two potential security implications:

- Fever of nearly 1 C (1-2 degrees Fahrenheit) could be masked by applying, to the human forehead, common cosmetics that contain solid particles.
- Fever of 1-2 C (almost 2-4 degrees Fahrenheit) could be masked temporarily, for up to 20-30 min, by applying common cosmetics to the human forehead.

## Data Availability

Data is available upon request from the authors.

## ACKNOWLEDGMENTS

This work was supported by the taxpayers of South Korea through the Institute for Basic Science, project code IBS-R020-D1.

## Notes

### Competing Interest Statement

The authors have declared no competing interest.

